# Risk factors for SARS-CoV-2 infection and hospitalisation in children and adolescents in Norway: A nationwide population-based study

**DOI:** 10.1101/2021.07.01.21259887

**Authors:** Ketil Størdal, Paz Lopez-Doriga Ruiz, Margrethe Greve-Isdahl, Pål Surén, Per Kristian Knudsen, Hanne Løvdal Gulseth, German Tapia

## Abstract

**Objective:** To determine risk factors for SARS-CoV-2 infection and hospitalisation among children and adolescents.

**Design:** Nationwide, population-based cohort study. Setting: Norway from 1 March 2020 to 31 April 2021. Participants: All Norwegian residents <18 years of age.

**Main outcome measures:** Population-based health care and population registries were used to study risk factors for SARS-CoV-2 infection, including socioeconomic factors, country of origin, and pre-existing chronic comorbidities. All residents were followed until age 18, emigration, death, or end of follow-up. Hazard ratios (HR) estimated by Cox regression models were adjusted for testing frequency. Further, risk factors for admission to the hospital among the infected were investigated.

**Results:** Of 1 182 796 residents, 22608 (1.9%) tested positive by polymerase chain reaction or lateral flow tests, of whom 107 (0.5%) were admitted to a hospital. Low family income (aHR 1.40, 95% confidence interval 1.36 to 1.46), crowded housing (1.35, 1.30 to 1.39), household size, age, and area of living were independent risk factors for infection. A non-Nordic country of origin was the strongest risk factor (aHR 2.37, 95% CI 2.30 to 2.49), whereas chronic comorbidity was not associated with the risk of infection. Chronic comorbidity was associated with hospitalisation (aHR 4.15, 2.63 to 6.56), in addition to age, whereas socioeconomic status and country of origin did not predict hospitalisation among those infected.

**Conclusions:** Socioeconomic factors, country of origin, and area of living were associated with the risk of SARS-CoV-2 infection. However, these factors did not predict hospitalisation among those infected. Chronic comorbidity was associated with the risk of admission but not with the overall risk of acquiring SARS-CoV-2.

**What is already known on this topic:** Hospital admissions rates for covid-19 among children and adolescents are low compared to adults. Admission rates to hospitals and intensive care units for covid-19 have been higher in minority groups and in children and adolescents with chronic comorbidity.

Whether underlying differences in susceptibility for severe disease or the incidence of infections are driving these associations have not been investigated.

**What this study adds:** Low family income, crowded housing and household size, and country of origin outside the Nordic countries were associated with increased risk of infection with SARS-CoV-2.

None of these factors, but chronic comorbidities, were associated with the risk of hospital admission among those infected.

## Introduction

The risk of severe covid-19 increases with age, as shown by the proportion of hospitalisation and death by age categories.^1^ A lower prevalence of antibodies in young children demonstrated in serology studies suggests that not only the risk of severe disease but also the risk of infection is lower in the youngest age groups.^2-4^

Exposure to infected household members is the primary source of infection,^5^ in addition to exposure from other contacts, including peers in day-care/school and leisure activities.^6^ Little is known regarding socioeconomic risk factors, such as household crowding, household size, and family income, for infection in children and adolescents.^5^ Country of origin has been associated with an increased risk of hospitalisation, but it is unclear whether this is related to socioeconomic factors or increased susceptibility to the virus.^7-10^

Chronic conditions that affect children may increase the risk of infection due to exposure through health and care services, particularly for children living in special care with many close adult contacts. Chronic conditions may also increase the severity of covid-19. In a European multicentre study, 25% of those included in a predominantly hospital-based sample had underlying medical conditions.^11^ Chronic conditions were associated with an increased risk of admission to an intensive care unit (ICU). In the United States, the prevalence of complex morbidity or neuromuscular disease was high among children admitted to ICU care.^12^ However, data on the baseline occurrence of these conditions in the general child population were not available, so the risk for hospitalisation associated with comorbidities could not be determined. A recent systematic review summarised the findings regarding comorbidities in severe covid-19,^13^ but studies tend to capture emergency hospital visits and inpatients and lack data on the vast majority who are infected without hospital encounters. Improved knowledge of the risk factors for infection and hospitalisation is relevant for mitigation and future vaccine strategies.

In this nationwide study covering the first 14 months of the pandemic, the aim was to determine risk factors for covid-19 in the population <18 years and to study risk factors for hospitalisation in those infected with SARS-CoV-2.

## Material and methods

We investigated the epidemiology of SARS-CoV-2 infections in a nationwide, population-based study. In an open cohort, inhabitants <18 years living in Norway at any time from 1^st^ March 2020 to 30^th^ April 2021 were included. The end of follow-up was 30^th^ April 2021, age 18 years, or death, whichever occurred first.

Individual-level data was available from the BEREDT C19 registry, developed specifically for emergency preparedness to provide knowledge on the spread of the SARS-CoV-2 virus.^14^ In the registry, the unique national identification number given to all citizens upon birth or immigration was used to link vital sources of information (Fig. 1):

**Fig. 1:**
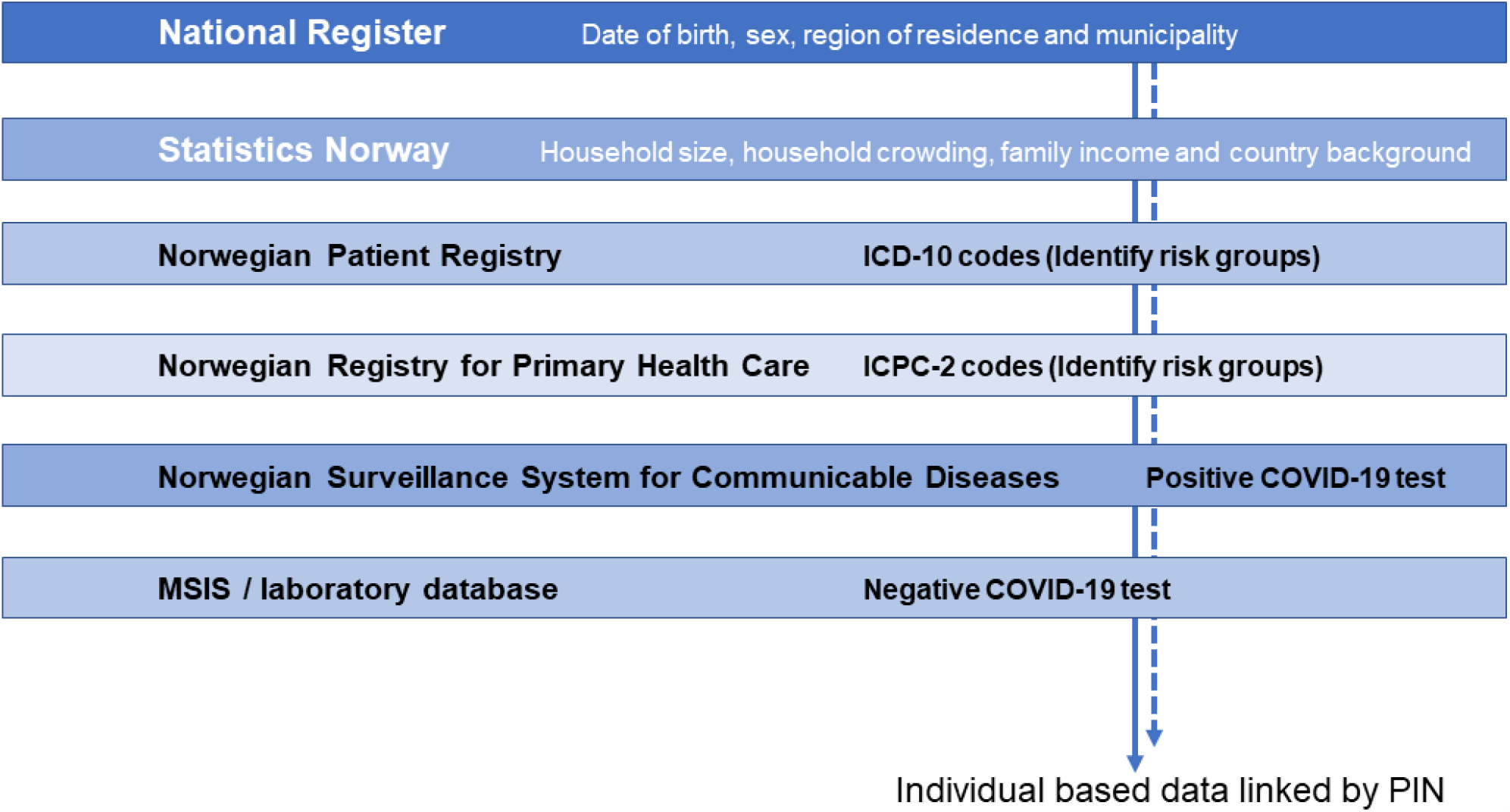
Flow of data and data sources for BEREDT C19.

- The National Population Registry includes information on date of birth, sex, municipality, and geographical region (South-East, West, Central, North).
- Statistics Norway (SSB) provides data on socioeconomic factors: household size, household crowding, family income, and country of origin.
- The Norwegian Patient Registry (NPR) is an administrative database that contains data on the activity at all publicly funded hospitals and clinics, including International Classification of Diseases (ICD-10) codes. Reporting to the NPR is mandatory and forms the basis for government reimbursements to specialist health services. The list of ICD-10 codes and their groups that were relevant for this study are provided in Fig. 2.
- The Norwegian Registry for Primary Health Care (NRPHC) covers claims for reimbursement from primary health service providers to the state. For this study, the International Classification for Primary Care (ICPC-2) code for asthma (R96) was used to capture milder forms of asthma not receiving specialist care. The other chronic conditions in our study were cared for at the specialist level; thus, further ICPC-2 codes were not used.
- The Norwegian Surveillance System for Communicable Diseases (MSIS) includes results of all positive and negative polymerase chain reaction (PCR) tests and rapid antigen tests for SARS-CoV-2. The dates of testing and test results are legally required to be reported from all laboratories to the MSIS. Some negative results could be missing before 1^st^ April 2020, but all positive results are included. Serology results were not available, except for in suspected cases of multisystem inflammatory syndrome (MIS-C).

**Fig. 2:**
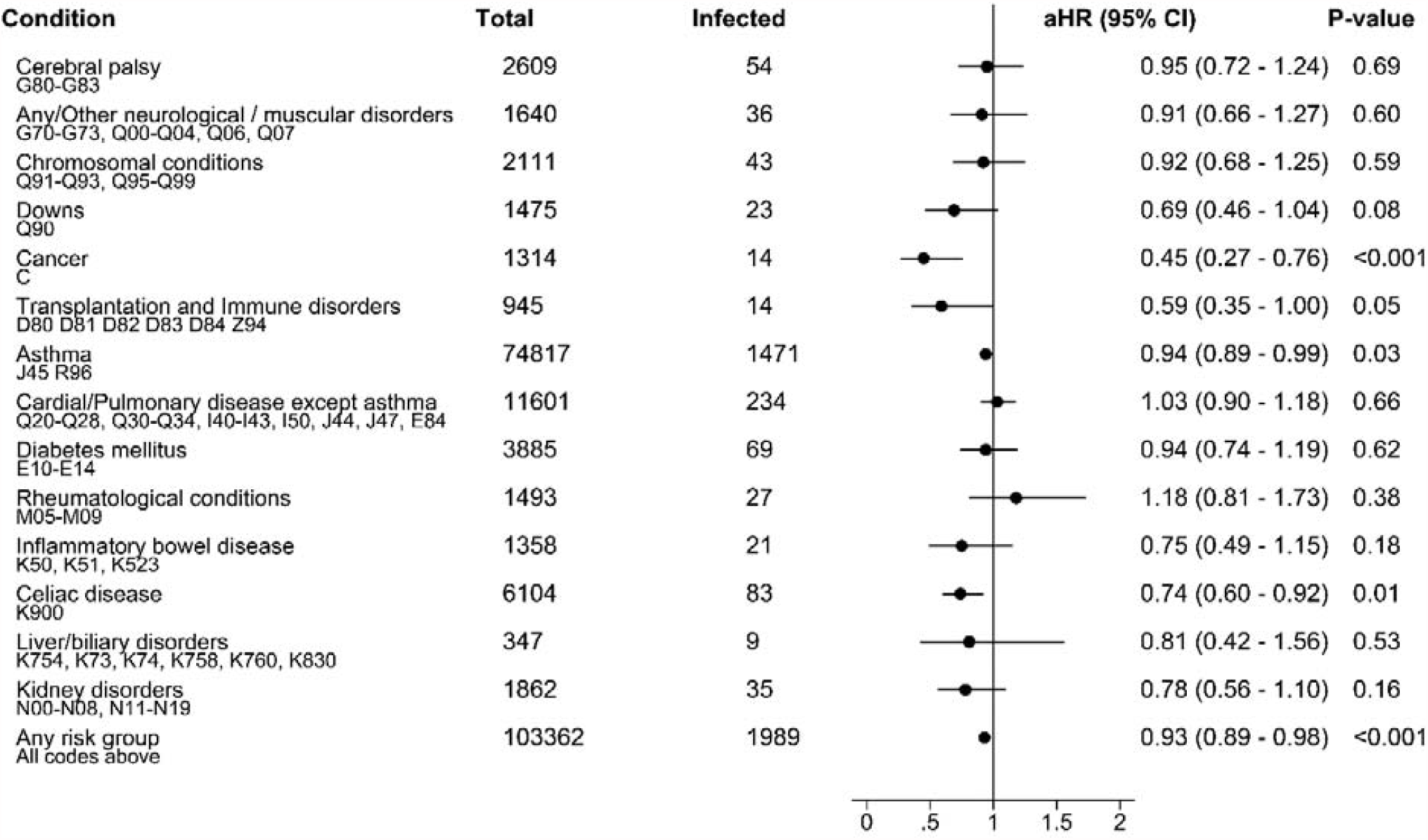
Pre-existing chronic condition in the population and in those infected with SARS-CoV-2 and adjusted hazard ratio for infection^*^. ^*^Adjusted for age, sex, region, municipality size, household size, household crowding, low family income, testing frequency and country of origin.

An institutional review board was conducted. The Regional Ethics Committee of South-East Norway confirmed (4^th^ June 2020, #153204) that an external ethical review board was not required for the use of BEREDT C19.

### Main outcomes

The main outcome was any infection by SARS-CoV-2, as confirmed by a PCR test or lateral flow (rapid test). To capture severe covid-19, hospitalisation (admission >5 hour duration) with a primary or secondary diagnosis of covid-19 (U07.1) and/or MIS-C (U10) was used. ICD-10 codes for admission with covid-19 were extracted to classify the main presentation (MIS-C, respiratory, gastrointestinal, other symptoms, and non-related diagnoses).

### Exposures

To characterise risk factors for infection and hospitalisation, we included sex and age (at time of a positive test, or at study entry) categorised into age groups (Table 1). The region of living (South-East vs West/Central/North) and size of the municipality (</≥50,000 residents) were further studied. Low family income was defined according to official statistics as <60% of the median income for Norway (last three years) for the family, weighted by the number of family members. Household crowding was defined by Statistics Norway as having fewer than one living room <u>and</u> <20 m^2^ of living space per household member. The size of the household was categorised as shown in Table 1. To study the potential impact of country of origin, we classified by the individuals’, maternal/paternal, and grandparents’ country of birth. Chronic comorbidities were grouped according to diagnostic codes, as shown in Fig. 2.

**Table 1:** Characteristics of Norwegian residents <18 years infected vs non-infected with SARS-CoV-2 from 1^st^ of March 2020 – 30^th^ of April 2021.

|  | No SARS-CoV-2 recorded<br>n=1 160 188 | SARS-CoV-2 positive<br>n=22 608<br>(1.91%) | Hazard ratio<br>(95% CI) | Adjusted<br>hazard ratio<br>(95% CI)<sup>§</sup> | p-value |
| --- | --- | --- | --- | --- | --- |
| **Age, n (%)<sup>\*</sup>** |  |  |  |  |  |
| ≤5 years | 407 417 (35.1) | 5 805 (25.7) | Ref. | Ref. |  |
| 6-11 years | 379 041 (32.7) | 7 842 (34.7) | 1.40 (1.35 - 1.45) | 1.01 (0.97 - 1.05) | 0.58 |
| 12-17 years | 373 730 (32.2) | 8 961 (39.6) | 1.91 (1.85 - 1.98) | 1.15 (1.11 - 1.19) | <0.001 |
| **Sex, n (%)** |  |  |  |  |  |
| Boys | 595 170 (51.3) | 11 692 (51.7) | Ref. | Ref. |  |
| Girls | 565 018 (48.7) | 10 916 (48.3) | 0.98 (0.96 - 1.01) | 0.99 (0.96 - 1.02) | 0.43 |
| **Geographical region, n (%)** |  |  |  |  |  |
| South-East | 642 408 (55.5) | 18 588 (82.3) | Ref. | Ref. |  |
| West/Central/<br>North | 515 372 (45.5) | 3 996 (17.7) | 0.27 (0.26 - 0.28) | 0.45 (0.43 - 0.46) | <0.001 |
| **Municipality size, n (%)<sup>†</sup>** |  |  |  |  |  |
| <50,000 | 626 963 (54.0) | 7 754 (34.3) | Ref. | Ref. |  |
| ≥50,000 | 530 817 (45.8) | 14 830 (65.6) | 2.24 (2.18 - 2.30) | 1.43 (1.39 - 1.48) | <0.001 |
| **Household size, n (%)<sup>†</sup>** |  |  |  |  |  |
| ≤2 | 87 245 (7.5) | 1 324 (5.9) | Ref. | Ref. |  |
| 3 | 229 844 (19.8) | 3 393 (15.0) | 0.96 (0.90 - 1.02) | 1.04 (0.97 - 1.10) | 0.27 |
| 4 | 452 399 (39.0) | 7 830 (34.6) | 1.10 (1.04 - 1.17) | 1.14 (1.07 - 1.20) | <0.001 |
| 5 | 249 458 (21.5) | 5 165 (22.8) | 1.32 (1.24 - 1.40) | 1.28 (1.20 - 1.36) | <0.001 |
| ≥6 | 95 466 (8.2) | 4 217 (18.7) | 2.80 (2.63 - 2.98) | 1.78 (1.66 - 1.90) | <0.001 |
| **Overcrowded living condition<sup>†</sup>** |  |  |  |  |  |
| n (%) | 204 042 (17.6) | 8 396 (37.1) | 2.70 (2.63 - 2.78) | 1.35 (1.30 - 1.39) | <0.001 |
| **Low family income<sup>†</sup>** |  |  |  |  |  |
| n (%) | 139 889 (12.1) | 6 108 (27.0) | 2.63 (2.55 - 2.70) | 1.40 (1.36 - 1.46) | <0.001 |
| **Country of origin, (n, %)** |  |  |  |  |  |
| Nordic countries<sup>‡</sup> | 731 628 (63.1) | 7 863 (34.8) | Ref. | Ref. |  |
| Europe | 159 783 (13.8) | 4 091 (18.1) | 2.33 (2.24 - 2.42) | 2.12 (2.04 - 2.21) | <0.001 |
| North America<br>and Oceania | 27 770 (2.4) | 346 (1.5) | 1.15 (1.03 - 1.28) | 1.02 (0.91 - 1.13) | 0.76 |
| Latin America | 20 794 (1.8) | 384 (1.7) | 1.69 (1.52 - 1.87) | 1.44 (1.30 - 1.60) | <0.001 |
| Middle East and<br>North Africa | 29 450 (2.5) | 2 093 (9.3) | 6.35 (6.05 - 6.66) | 3.61 (3.42 - 3.80) | <0.001 |
| Africa | 50 620 (4.4) | 3 105 (13.7) | 5.50 (5.28 - 5.74) | 3.43 (3.27 - 3.59) | <0.001 |
| Asia | 101 815 (8.8) | 4 329 (19.1) | 3.86 (3.72 - 4.01) | 2.77 (2.66 - 2.88) | <0.001 |

<sup>\*</sup> Age at inclusion in study. Children that died at birth (n = 17) and born 30 April 2021 were excluded. Five children that were SARS-CoV-2 positive at birth were included.
<sup>†</sup> Missing region and municipality size n=2 408, household size/low family income n=45 776, overcrowded living condition n=64 729, country of origin n=38 328. The youngest children (born in 2020 and 2021) are predominantly missing in socioeconomic covariates and country of origin.
<sup>‡</sup> Nordic countries: Norway, Sweden, Finland, Denmark, Iceland and Faroe islands.
<sup>§</sup> adjusted for all covariates in the table and additionally for testing frequency.

### Statistics

We performed summary statistics for categorical variables, summarising absolute numbers and percentages. Cox regression models were further used to estimate hazard ratios (HRs) for confirmed infection or hospitalisation before the end of follow-up. Proportional hazards were assessed by log-log plots of survival. In the main analysis, we included the aforementioned sets of *a priori* selected covariates in multivariable models. To account for differences in testing practices and policies, the frequency of testing was adjusted for, with a maximum of one test recorded per week to account for clustered tests due to outbreak investigations.

The frequency of hospitalisation was low among those who were infected. In the analysis of hospitalisation risk, we dichotomized the country of origin and chronic comorbidities. We further explored hospitalisation for primary infection and with MIS-C separately to assess whether comorbidities were risk factors specific to these outcomes. In pre-planned sensitivity analyses, we stratified our analyses for 2020 vs. 2021 because of the introduction of the alpha virus variant in January 2021 and increased test activity in 2021, which may have impacted detection rates. Analyses were further stratified by counties which had a higher spread of infection (Oslo and the surrounding county Viken) compared to the nine other counties in Norway.

### Patient and public involvement

Patients were not involved in setting the research question or the outcome measures. No patients were asked to advise on interpretation or writing up of results. We will disseminate the results of the research to the general public.

## Results

During the observation period of 14 months, 22608 of 1 182 796 inhabitants <18 years had a positive test for SARS-CoV-2, yielding a cumulative incidence of 19.1/1000 (Table 1). The incidence increased by age category but did not differ by sex (Fig. 3). After adjustments, the risk of infection remained higher from 12-17 years compared to the reference category of ≤5 year (Table 1).

**Fig. 3:**
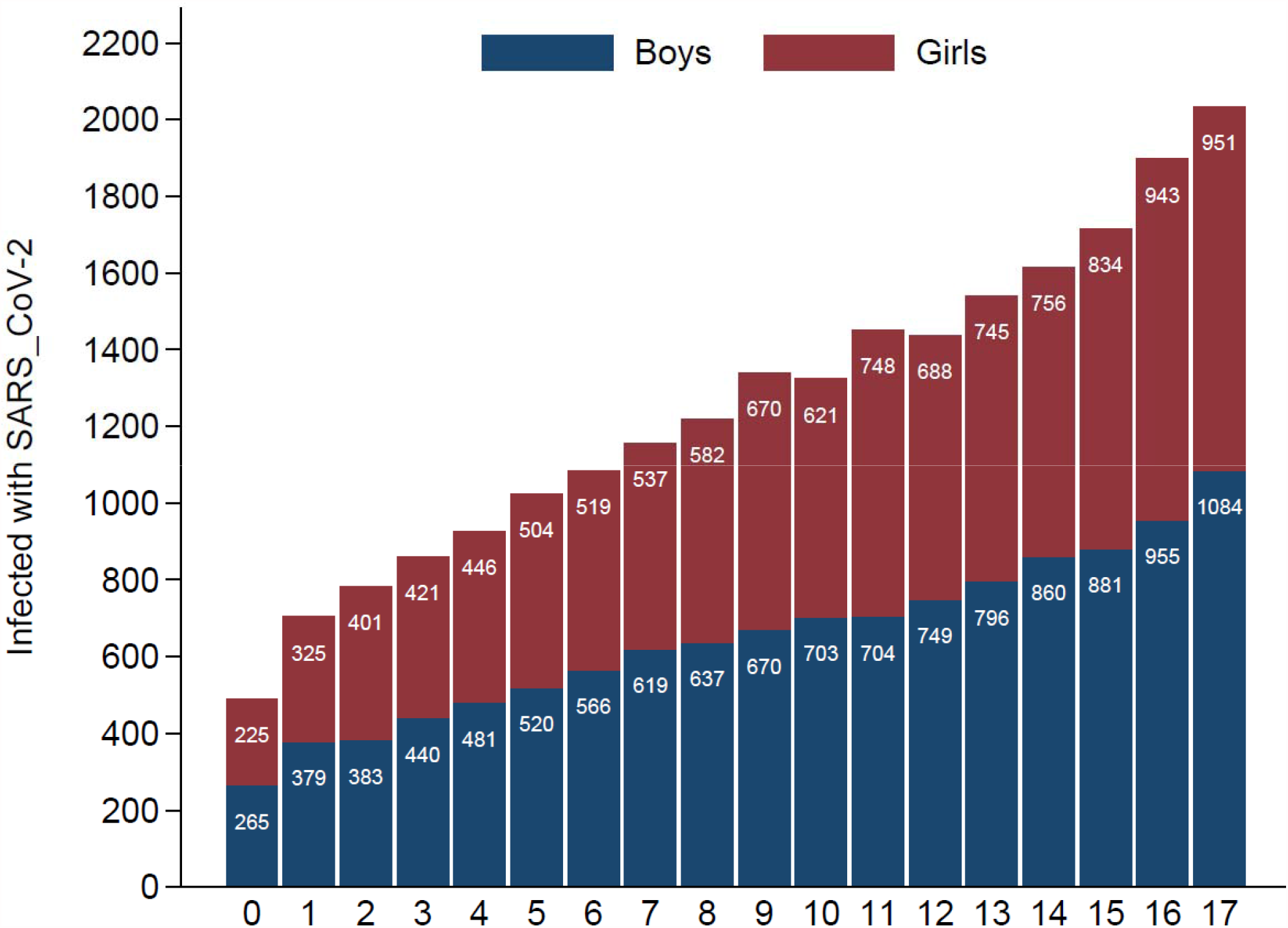
Age at time of positive test and sex of infected with SARS-CoV-2 (cumulative cases 1^st^ of March 2020 to 30^th^ of April 2021)

Of those infected, 107/22608 (4.7/1000) were admitted to a hospital. MIS-C was recorded in 22, yielding an incidence of 1.0/1000 in those with known infection. Of the remaining 85 subjects who were hospitalised, 32 had respiratory codes, seven had gastroenterological and seven respiratory/gastroenterological codes, and 20 had other symptom codes. Nineteen were admitted for reasons other than covid-19, of whom five were infected from birth. The median hospital stay length was two days, and 89% were discharged within 7 days. Admission to intensive care (n=11) and death (n=2) were rare events, and these outcomes were not studied further.

### Socioeconomic and demographic risk factors for infection

The incidence of infection was highest in the South-East region, and those living in municipalities with >50000 inhabitants had a significantly higher risk compared with smaller municipalities (adjusted HR [aHR] 1.43, 95% CI 1.39 to 1.48, Table 1). Living in households with ≥4 members compared with smaller households was associated with an increased risk, and particularly increased with ≥6 in the household (aHR 1.78, 95% CI 1.66 to 1.90, Table 1). Household crowding and low family income were independent and significant predictors for the risk of infection (p<0.001, Table 1).

Country origin outside Nordic countries was associated with an increased risk (aHR 2.37, 95% CI 2.30 to 2.49). The risk estimates were highest for residents with family backgrounds from Africa, Asia, and the Middle East/North Africa, whereas the estimates for North America/Oceania were similar as those observed for Nordic countries (Table 1). Age category, region of living and municipality size, low income, household crowding and size, and country of origin were independent predictors for infection with SARS-CoV-2 in the adjusted model (Table 1).

### Chronic conditions as risk factors for infection

Of the total population, 103 362 (8.7%) had diagnoses of chronic conditions, as listed in Fig. 2. Overall, there was a slightly lower risk of being infected with SARS-CoV-2 in children and adolescents with chronic conditions (aHR 0.93, 95% CI 0.89 to 0.98, Fig. 2). None of the groups of chronic comorbidities had a significantly increased risk in unadjusted or adjusted analyses (Fig. 2). Notably, we found a significantly lower risk of infection for those with cancer and the prevalent immune-mediated conditions of asthma and coeliac disease (Fig. 2).

### Risk factors for hospitalisation

Among those infected with SARS-CoV-2, the risk of hospitalisation was lowest for the age group 6-11 years (Table 2). Sociodemographic factors were not associated with the risk of hospital admission. A non-Nordic country of origin was not a risk factor for hospitalisation among those infected (aHR 1.42, 95% CI 0.85 to 2.35, Table 2). Among those admitted to a hospital, 27 of 107 (25%) had a chronic comorbidity, which was a strong risk factor for hospital admission (aHR 4.15, 95% CI 2.63 to 6.56). Asthma and other chronic cardiopulmonary conditions were the most prevalent, and the latter group was particularly associated with hospitalisation (Table 2). When excluding those admitted with a diagnosis of MIS-C or admission unrelated to covid-19, the risk estimates for chronic comorbidities were not substantially changed (aHR 4.47, 95% CI 2.51 to 7.95). Chronic comorbidity was also associated with increased risk of MIS-C (aHR 3.58, 95% CI 1.31 to 9.80).

**Table 2:** Risk for hospital admission among residents <18 years infected with SARS-CoV-2 (n=22 608).

| Exposure | Hospitalized, n (%) |  | Hazard ratio<br>(95% CI) | Adjusted hazard<br>ratio (95% CI)<sup>*</sup> | p-value |
| --- | --- | --- | --- | --- | --- |
|  | Yes<br>(n=107) | No<br>(22 501) |  |  |  |
| Age category, (n,%)<sup>†</sup> |  |  |  |  |  |
| ≤5 years | 49 (45.8) | 5 756 (26.0) | Ref. | Ref. |  |
| 6-11 years | 15 (14.0) | 7 827 (34.8) | 0.25 (0.14 - 0.44) | 0.34 (0.18 - 0.64) | <0.001 |
| 12-17 years | 43 (40.2) | 8 918 (39.6) | 0.64 (0.42 - 0.97) | 0.89 (0.56 - 1.42) | 0.62 |
| Sex, n (%) |  |  |  |  |  |
| Boys | 54 (50.5) | 11 683 (51.7) | Ref. | Ref. |  |
| Girls | 53 (49.5) | 10 863 (48.3) | 1.03 (0.70 - 1.52) | 1.03 (0.68 - 1.57) | 0.56 |
| Region, n (%) |  |  |  |  |  |
| South-East | 79(73.8) | 18 509 (82.3) | Ref. | Ref. |  |
| Others | 27 (26.2) | 3 992 (17.7) | 1.62 (1.04 - 2.54) | 1.81 (1.12 - 2.91) | 0.03 |
| Municipality size, n (%)<sup>‡</sup> |  |  |  |  |  |
| <50,000 | 36 (33.6) | 7 718 (34.3) | Ref | Ref |  |
| ≥50,000 | 70 (65.4) | 14 760 (65.6) | 0.99 (0.65 - 1.49) | 0.90 (0.58 - 1.40) | 0.63 |
| Household size, (n,%)<sup>‡</sup> |  |  |  |  |  |
| ≤2 | 9 (8.4) | 1 315 (5.8) | Ref | Ref |  |
| 3 | 19 (17.8) | 3 374 (15.0) | 1.23 (0.49 - 3.07) | 1.17 (0.46 - 2.95) | 0.74 |
| 4 | 34 (31.8) | 7 796 (34.6) | 0.95 (0.40 - 2.25) | 1.08 (0.44 - 2.61) | 0.87 |
| 5 | 19 (17.8) | 5 146 (22.9) | 0.80 (0.32 - 2.01) | 0.86 (0.33 - 2.25) | 0.76 |
| ≥6 | 14 (13.1) | 4 203 (18.7) | 0.72 (0.28 - 1.88) | 0.74 (0.26 - 2.09) | 0.57 |
| Low family income, (n, %)<sup>‡</sup> |  |  |  |  |  |
|  | 31 (29.0) | 6 077 (27.0) | 1.25 (0.81 - 1.94) | 1.43 (0.87 - 2.37) | 0.16 |
| Overcrowded living conditions, (n, %)<sup>‡</sup> |  |  |  |  |  |
|  | 33 (30.8) | 8 363 (37.2) | 0.87 (0.57 - 1.34) | 0.87 (0.51 - 1.49) | 0.61 |
| Country background, (n, %) |  |  |  |  |  |
| Nordic countries | 29 (27.1) | 7 834 (34.8) | Ref | Ref |  |
| Non-Nordic | 78 (72.9) | 14 667 (65.2) | 1.34 (0.87 - 2.07) | 1.42 (0.85 - 2.35) | 0.18 |
| Chronic condition, (n, %) |  |  |  |  |  |
| No | 80 (74.8) | 20 539 (91.3) | Ref | Ref |  |
| Any | 27 (25.2) | 1 962 (8.7) | 3.75 (2.41 - 5.82) | 4.15 (2.63 - 6.56) | <0.001 |
| Asthma<sup>§</sup> | 12 (11.2) | 1 459 (6.5) | 1.91 (1.05 - 3.49) | 2.18 (1.19 - 4.02) | <0.001 |
| Chronic cardiopulmonary<br>except asthma<sup>‡</sup> | 9 (8.4) | 225 (1.0) | 9.47 (4.78 - 18.8) | 9.62 (4.76 – 19.5) | <0.001 |
| Cerebral palsy/other<br>neuromuscular/ Downs/<br>other chromosomal | n < 5 | 147 (0.7) | 4.57 (1.45 -14.4) | 3.57 (0.88 – 14.6) | 0.08 |
| Cancer, transplantation<br>and immunodeficiencies<sup>§</sup> | n < 5 | 24 (0.1) | 34.5 (12.7 - 93.8) | 35.0 (12.7 – 96.8) | <0.001 |
| Liver/kidney disorders<sup>§</sup> | n < 5 | 40 (0.2) | 22.5 (8.28 - 61.2) | 24.3 (8.9 – 67.0) | <0.001 |
| Autoimmune disorders<sup>§</sup> | 0 | 0 | NA | NA | NA |

\* Adjusted for all other covariates in the table.
<sup>†</sup> Age at admission or positive test for SARS-CoV-2.<sup>‡</sup> Missing municipality size n=1, household size, low family income and overcrowded living conditions n=12<sup>§</sup> These sub-analyses were run separately without adjustment for other comorbidities.<sup>||</sup> Due to privacy guidelines we are unable to show exact numbers in cells with fewer than five individuals.

Stratified analyses for risk factors for infection by time period showed a difference for age categories, with an increased risk in age groups 6-11 and 12-17 in the first period which was not present in the last period (supplementary Table 1). Analyses stratified by geographical area did not indicate substantial differences between Oslo/Viken vs. other counties (supplementary Table 2).

## Discussion

The main risk factors for infection with SARS-CoV-2 among children and adolescents in the present study were socioeconomic determinants and country of origin, in addition to geographic region and municipality size. Hospitalisation for covid-19 was very uncommon, but premorbid chronic conditions and young age were associated with increased risk. The country of origin and socioeconomic factors were not associated with the risk for hospitalisation among those infected.

### Strengths and limitations of study

Among several strengths of the current study, sample size provided by the linkage of nationwide registers and avoidance of a selection bias, which is often encountered in hospital-based studies, are prominent. To the best of our knowledge, this is the first large study to determine socioeconomic characteristics and country of origin as risk factors of SARS-CoV2-infection across the range of severity in children and adolescents. The coverage of this nationwide study was high, likely capturing the majority of all infected, as suggested by seroprevalence studies indicating that the majority of cases in our country were detected by PCR.^15^ However, the availability of testing was limited during the first months of the pandemic. This may have resulted in a higher proportion of undetected cases during the first period, particularly among children and adolescents.

Furthermore, the linkage to national diagnosis registers provides trustable detection of relevant chronic comorbidities. A recording of overweight/obesity was not available, and this factor has also been associated with covid-19 severity in children and adolescents.^9, 10, 13^ Risk factors for disease severity may be biased if chronic disease were part of the test criteria, which to some extent occurred during the early phases of the pandemic. Comorbidity and young age would likely lower the threshold for hospital admission, potentially inflating the observed associations.

Country of origin was the strongest predictor for the risk of infection. Notably, these risk estimates remained highly significant when adjusting for socioeconomic factors. This suggests that country of origin and socioeconomic status were independent factors for the distribution of SARS-CoV-2 in our population. A report from the US showed a higher risk of infection in non-white ethnic groups, but did not account for socioeconomic factors.^16^ How such factors confound associations with country of origin in societies with welfare systems that are different from the publicly funded health care system in Scandinavia should be studied further.

The higher rates of SARS-CoV-2 in non-Nordic groups has also been observed for adults in Norway.^17^ Because family contacts are the main source of infection, a similar difference by country of origin is expected among children and adolescents. Factors related to a higher prevalence in these families are higher occupational exposure and contact with high-endemic areas by travel or visitors.

Communication of the implemented strict infection control measures may not reach certain groups, and cultural differences in interpersonal contact may increase the vulnerability to infectious diseases.

### Comparison with other studies

Country of origin was not a risk factor for hospitalisation in our study. This is important, as other studies have also raised concern for such an association in children, and this finding was in contrast to most previous studies among hospital-based cohorts of children and adolescents.^7-10^ However, these studies did not provide data that separated the risk of infection from hospitalisation. The current data suggests that a skewed distribution of country of origin among hospitalised children was driven by differences in the spread of infection in society and not by susceptibility to severe disease, in line with a recent US study on risk factors for severe covid-19.^18^ In adults, genetic susceptibility clearly influences the severity of covid-19 infection,^19, 20^ which is attributed to gene variants that are differentially distributed across ethnic groups.^21^ Such a genetic susceptibility remains to be proven for severe covid-19 disease among children and adolescents.

The linkage to nationwide registers allowed us to study pre-existing comorbidities as a risk factor for infection and hospital admission. This differs from previous studies reporting whether children and adolescents that were admitted to a hospital with a chronic comorbidity had an increased risk of ICU transfer or death.^7-10^ Because a very low percentage of children and adolescents required admission (0.47%), our findings provide new information on risk factors for infection regardless of severity. None of the chronic comorbidities were associated with an increased risk of infection. The national policy during the pandemic was to keep schools and kindergartens open with infection control measures, and only children with severe chronic comorbidities were advised to stay home. However, stricter infection control in vulnerable groups may have led to a somewhat lower risk of infection. Vaccination of adolescents with comorbidities (≥16 years age) started in March 2021 and is unlikely to affect our findings because <700 had received the vaccine by the end of April 2021.

The excess risk for hospitalisation for children with any comorbidity of HR 4.15 was in line with a large study from the CDC COVID-19-NET, including over 250 centres reporting an adjusted odds ratio (aOR) of 3.55 (95% CI 3.14 to 4.01)^7^. However, 88% in the CDC study were excluded due to missing data, which could bias the associations. Furthermore, chronic comorbidity was a risk factor for hospital admission among 454 children from Colorado (OR 2.73), of whom 15% were admitted due to symptomatic infection.^22^

Comorbidity was a risk factor for ICU admission or death among hospitalised children and adolescents in a multi-centre study from the first wave in the UK, in which 42% of hospitalised children had at least one comorbidity and all who died (6/627) had a severe comorbidity.^9^ Similarly, a European study and an American multi-site study found increased risk for ICU admission in those with a chronic comorbidity (aOR 3.27 in both studies).^11, 18^ A complex comorbidity was not a significant risk factor for severity (aOR 1.51, 95% CI 0.51 to 4.42) in a US multi-centre study, although four of seven deaths occurred in such children.^10^ Comorbidities were recorded in 65% of those with severe, compared to 25% of non-severe, infections in a large French hospital-based study (aOR 2.9, 95% CI 0.9 to 9.9).^23^ MIS-C differed from other admissions characterised by a low occurrence of pre-existing comorbidity in the studies of hospitalised cases from the US, UK, and France.^9, 10, 23^ This contrasts our findings, however our study included few MIS-C cases and should be interpreted cautiously.

Asthma was the most prevalent comorbidity in our study. Children with asthma had a slightly lower risk of infection and a somewhat increased risk of hospital admission compared to children without asthma. Current asthma has been suggested to be negatively associated with the risk of hospitalisation for Covid-19 in children^24,25^ and was not associated with hospitalisation in a study including both children and adults.^26^ Similar to our study, other chronic cardiopulmonary conditions have been reported as risk factors for hospital admission and/or ICU transfer.^9, 11, 23^ The current study had a limited number of hospitalised children, precluding precise estimates for less frequent comorbidities. Neurological and congenital disorders, malignancy, immunocompromise, and gastrointestinal comorbidities have, to a varying extent, been associated with hospitalisation or ICU admission.^11, 13, 22,23^

Currently, the vaccine roll-out and discussions about whether children and adolescents should be vaccinated are ongoing. Groups of children and adolescents at risk of a severe course are important to identify, and determinants of a higher risk of infection are relevant if a targeted vaccination strategy is considered. Mitigation strategies in addition to vaccination could also focus on groups with an increased incidence of SARS-CoV-2 or severe complications. Socioeconomic disparities were recently demonstrated in Brazil^27^ but should be further studied in high-income countries. Through public health efforts, particularly with testing, contact tracing, quarantine, and isolation, Norway succeeded in limiting the spread of SARS-CoV-2. A lower incidence of SARS-CoV-2 compared to most other countries may reduce the generalisability of our findings.

## Conclusion

The results from the current study provides novel data on the socioeconomic determinants of infection. A strong association with country of origin suggests that non-pharmaceutical and pharmaceutical interventions targeted to minority groups of children and adolescents could mitigate further disease. Chronic comorbidity was associated with the risk of admission but not with the overall risk of acquiring SARS-CoV-2.

## Data Availability

No additional data are available.

## Copyright

*The Corresponding Author has the right to grant on behalf of all authors and does grant on behalf of all authors, an exclusive licence (or non-exclusive for government employees) on a worldwide basis to the BMJ Publishing Group Ltd to permit this article (if accepted) to be published in BMJ editions and any other BMJPGL products and sublicenses such use and exploit all subsidiary rights, as set out in our licence*.

## Contributors

KS and GT coordinated the study, wrote the analysis plans and had the primary responsibility for writing the paper. GT and PLDR did the statistical analysis and reviewed and commented on drafts. HLG supervised the study, interpreted the data and reviewed and commented on all drafts. MGI, PS and PKK reviewed the drafts and contributed in the interpretation of the findings.

The corresponding author attests that all listed authors meet authorship criteria and that no others meeting the criteria have been omitted.

## Data sharing

No additional data available.

## Transparency

The lead author (KS) affirms that the manuscript is an honest, accurate, and transparent account of the study being reported; that no important aspects of the study have been omitted; and that any discrepancies from the study as planned have been explained.

