## Supplemental tables for "Risk factors for SARS-CoV-2 infection and hospitalisation in children and adolescents in Norway: A nationwide population-based study"

**Supplementary Table 1:** Stratified analysis comparing risk factors for SARS-CoV-2 infection the first and the second time period among children and adolescents in Norway.

|  | **Prior to 1^st^ January 2021** | **After 1^st^ January**  **2021** |
| --- | --- | --- |
| **Exposure** | **aHR**^*^**, 95% CI** | **aHR**^*^**, 95% CI** |
| **Age category** |  |  |
| ≤5 years | Ref. | Ref. |
| 6-11 years | 1.30 (1.21 - 1.39) | 0.92 (0.88 - 0.96) |
| 12-17 years | 1.77 (1.65 - 1.88) | 0.94 (0.90 - 0.98) |
| **Female sex** | 0.98 (0.94 - 1.03) | 0.99 (0.96 - 1.02) |
| **Country of origin** |  |  |
| Nordic countries | Ref. | Ref. |
| Europe | 1.96 (1.83 - 2.10) | 2.20 (2.09 - 2.30) |
| North America and Oceania | 0.97 (0.80 - 1.17) | 1.04 (0.91 - 1.18) |
| Latin America | 1.30 (1.08 - 1.57) | 1.52 (1.34 - 1.72) |
| Middle East and North Africa | 3.32 (3.03 - 3.64) | 3.75 (3.52 - 3.99) |
| Africa | 3.79 (3.49 - 4.10) | 3.25 (3.07 - 3.45) |
| Asia | 2.54 (2.36 - 2.73) | 2.87 (2.74 - 3.02) |
| **Non South-East health region** | 0.59 (0.55 - 0.62) | 0.38 (0.36 - 0.40) |
| **Urban municipality** | 1.56 (1.48 - 1.64) | 1.37 (1.33 - 1.42) |
| **Household size** |  |  |
| ≤2 | Ref. | Ref. |
| 3 | 1.10 (0.98 - 1.24) | 1.01 (0.93 - 1.09) |
| 4 | 1.22 (1.09 - 1.35) | 1.10 (1.03 - 1.19) |
| 5 | 1.37 (1.22 - 1.53) | 1.24 (1.15 - 1.34) |
| ≥6 | 1.84 (1.63 - 2.07) | 1.75 (1.62 - 1.90) |
| **Low family income** | 1.46 (1.37 - 1.55) | 1.38 (1.32 - 1.44) |
| **Overcrowded living conditions** | 1.27 (1.19 - 1.34) | 1.39 (1.33 - 1.45) |
| **Chronic conditions** |  |  |
| Cerebral palsy | 0.86 (0.53 - 1.40) | 0.99 (0.72 - 1.37) |
| Any/Other neurological/muscular disorders | 1.23 (0.74 - 2.04) | 0.77 (0.49 - 1.19) |
| Chromosomal conditions | 1.68 (1.11 - 2.52) | 0.58 (0.37 - 0.93) |
| Downs syndrome | 0.60 (0.27 - 1.34) | 0.73 (0.46 - 1.18) |
| Cancer | 0.41 (0.15 - 1.08) | 0.47 (0.25 - 0.88) |
| Transplantation and immune disorders | 0.28 (0.07 - 1.11) | 0.73 (0.42 - 1.29) |
| Asthma | 0.92 (0.84 - 1.01) | 0.95 (0.89 - 1.01) |
| Chronic cardial or pulmonary disease except asthma | 1.24 (0.99 - 1.55) | 0.94 (0.80 - 1.11) |
| Diabetes mellitus | 0.80 (0.52 - 1.23) | 1.02 (0.77 - 1.36) |
| Rheumatological conditions | 1.34 (0.74 - 2.42) | 1.10 (0.67 - 1.80) |
| Inflammatory bowel disease | 0.78 (0.39 - 1.56) | 0.73 (0.43 - 1.26) |
| Celiac disease | 0.63 (0.42 - 0.94) | 0.80 (0.62 - 1.03) |
| Liver/biliary disorders | 0.27 (0.04 - 1.90) | 1.10 (0.55 - 2.20) |
| Kidney disorders | 0.74 (0.40 - 1.37) | 0.81 (0.54 - 1.22) |
| **Any risk condition** | 0.92 (0.84 - 0.99) | 0.94 (0.89 - 0.99) |

^*^adjusted for all covariates in the table and additionally for testing frequency

**Supplementary Table 2:** Stratified analysis comparing risk factors for SARS-CoV-2 infection in the two counties having the highest SARS-CoV-2 incidence with the other nine counties of Norway.

|  | **Oslo and Viken** | **Other parts of Norway** |
| --- | --- | --- |
| **Exposure** | **aHR**^*^**, 95% CI** | **aHR**^*^**, 95% CI** |
| **Age category** |  |  |
| ≤5 years | Ref. | Ref. |
| 6-11 years | 1.01 (0.96 - 1.07) | 1.01 (0.97 - 1.06) |
| 12-17 years | 1.17 (1.10 - 1.23) | 1.15 (1.10 - 1.20) |
| **Female sex** | 0.96 (0.92 - 1.00) | 1.01 (0.97 - 1.04) |
| **Country of origin** |  |  |
| Nordic countries | Ref. | Ref. |
| Europe | 2.00 (1.89 - 2.12) | 2.12 (2.01 - 2.24) |
| North America and Oceania | 1.08 (0.91 - 1.28) | 1.01 (0.88 - 1.16) |
| Latin America | 1.19 (0.99 - 1.43) | 1.58 (1.39 - 1.79) |
| Middle East and North Africa | 3.57 (3.30 - 3.86) | 3.51 (3.27 - 3.76) |
| Africa | 2.99 (2.76 - 3.24) | 3.64 (3.43 - 3.87) |
| Asia | 2.62 (2.47 - 2.79) | 2.78 (2.63 - 2.93) |
| **Urban municipality** | 1.17 (1.12 - 1.22) | 1.81 (1.74 - 1.89) |
| **Household size** |  |  |
| ≤2 | Ref. | Ref. |
| 3 | 1.23 (1.10 - 1.37) | 0.93 (0.85 - 1.00) |
| 4 | 1.32 (1.20 - 1.46) | 1.02 (0.94 - 1.09) |
| 5 | 1.49 (1.34 - 1.66) | 1.16 (1.07 - 1.25) |
| ≥6 | 2.14 (1.91 - 2.39) | 1.59 (1.46 - 1.73) |
| **Low family income** | 1.41 (1.34 - 1.50) | 1.44 (1.38 - 1.50) |
| **Overcrowded living conditions** | 1.26 (1.20 - 1.33) | 1.43 (1.37 - 1.49) |
| **Chronic conditions** |  |  |
| Cerebral palsy | 0.80 (0.50 - 1.27) | 1.05 (0.75 - 1.47) |
| Any/Other neurological/muscular disorders | 0.65 (0.36 - 1.18) | 1.13 (0.75 - 1.68) |
| Chromosomal conditions | 0.91 (0.55 - 1.50) | 0.96 (0.65 - 1.41) |
| Downs | 0.66 (0.34 - 1.26) | 0.72 (0.42 - 1.21) |
| Cancer | 0.34 (0.13 - 0.90) | 0.53 (0.28 - 0.98) |
| Transplantation and immune disorders | 0.53 (0.22 - 1.27) | 0.66 (0.34 - 1.27) |
| Asthma | 0.98 (0.90 - 1.06) | 0.91 (0.85 - 0.98) |
| Chronic cardial or pulmonary disease except asthma | 0.98 (0.79 - 1.21) | 1.06 (0.90 - 1.25) |
| Diabetes mellitus | 0.96 (0.66 - 1.41) | 0.95 (0.70 - 1.28) |
| Rheumatological conditions | 1.67 (1.00 - 2.76) | 0.87 (0.49 - 1.53) |
| Inflammatory bowel disease | 0.85 (0.46 - 1.58) | 0.68 (0.37 - 1.22) |
| Celiac disease | 0.83 (0.58 - 1.17) | 0.71 (0.54 - 0.94) |
| Liver/biliary disorders | 1.11 (0.42 - 2.95) | 0.68 (0.28 - 1.64) |
| Kidney disorders | 0.82 (0.50 - 1.36) | 0.75 (0.47 - 1.19) |
| **Any risk condition** | 0.96 (0.89 - 1.03) | 0.91 (0.86 - 0.97) |

^*^adjusted for all covariates in the table and additionally for testing frequency
